# Integrating Expert Knowledge into Large Language Models Improves Performance for Psychiatric Reasoning and Diagnosis

**DOI:** 10.1101/2025.07.19.25331840

**Authors:** Karthik V Sarma, Kaitlin E Hanss, Andrew J M Halls, Andrew Krystal, Daniel F Becker, Anne L Glowinski, Atul J Butte

**Affiliations:** Department of Psychiatry and Behavioral Sciences, University of California San Francisco, 675 18th Street, San Francisco, CA 94143; Bakar Computational Health Sciences Institute, University of California San Francisco, 550 16th Street, San Francisco, CA, 94143

**Author notes:** Corresponding author: Karthik V Sarma MD PhD, Department of Psychiatry and Behavioral Sciences University of California San Francisco, 675 18th Street, Box 3134 San Francisco, CA 94143. In memoriam (Atul J Butte). **Previous Presentation:** The authors appreciated the opportunity to present early partial components of this project at the annual meetings of the Northern California Psychiatric Society (conference abstract/poster, Mar 16, 2024), the American Medical Informatics Association (conference abstract/talk, Nov 12, 2024), the Technology in Psychiatry Summit (symposium abstract/talk, Dec 7, 2024), and the American College of Neuropsychopharmacology (conference abstract/poster, Dec 10, 2024). No text, tables or figures used in these presentations were reused for this work.

**Keywords:** diagnosis, DSM, artificial intelligence, generative artificial intelligence, psychiatry, reasoning, decision support

## Abstract

**Purpose and Methods:** The authors sought to evaluate the performance of common large language models (LLMs) in psychiatric diagnosis, and the impact of integrating expert-derived reasoning on their performance. Clinical case vignettes and associated diagnoses were retrieved from the DSM-5-TR Clinical Cases book. Diagnostic decision trees were retrieved from the DSM-5-TR Handbook of Differential Diagnosis and refined for LLM use. Three LLMs were prompted to provide diagnosis candidates for the vignettes either by directly prompting or using the decision trees. These candidates and diagnostic categories were compared against the correct diagnoses. The positive predictive value (PPV), sensitivity, and F_1_ statistic were used to measure performance.

**Principal Results:** When directly prompted to predict diagnoses, the best LLM by F_1_ statistic (gpt-4o) had sensitivity of 77.6% and PPV of 43.3%. When making use of the refined decision trees, PPV was significantly increased (65.3%) without a significant reduction in sensitivity (71.8%). Across all experiments, the use of the decision trees statistically significantly increased the PPV, significantly increased the F_1_ statistic in 5/6 experiments, and significantly reduced sensitivity only for the category-based evaluation in 2/3 experiments.

**Major Conclusions:** When used to predict psychiatric diagnoses from case vignettes, direct prompting of the LLMs yielded most true positive diagnoses but had significant overdiagnosis. Integrating expert-derived reasoning into the process using decision trees improved LLM performance, primarily by suppressing overdiagnosis with minimal negative impact on sensitivity. This suggests that the integration of clinical expert-derived reasoning could improve the performance of LLM-based tools in the behavioral health setting.

## 1. Introduction

The rapid advancement of artificial intelligence (AI)-based technologies over the course of the last decade has led to dramatic innovation in healthcare technology across a wide variety of functional areas and clinical domains. Stemming both from rapid advancements in processing hardware and theoretical computer science, AI methods have proven particularly applicable to healthcare, with recent major results and novel products in radiology (Sarma et al., 2021), ophthalmology (Lim et al., 2024; Nguyen et al., 2024), emergency medicine (Bains et al., 2024; Williams et al., 2024b, 2024a), and many other fields. One challenge in the use of AI-based approaches in behavioral health has been the challenge of working with unstructured text-based data, such as provider progress notes, nursing notes, and provider interviews. Such records are generally not standardized and exhibit significant variability even within a single setting.

Recent advances in generative AI technologies have led to the development of large language models (LLMs), such as OpenAI’s ChatGPT. LLMs, designed for use in natural language tasks, are text-to-text predictive models trained on very large corpora of unstructured text that have shown great promise in nature language processing and understanding. Within behavioral health, LLMs have been investigated for use in documentation review and creation (Tierney et al., 2024), analysis of clinical text for decision support in diagnosis and treatment (Galatzer-Levy et al., 2023; So et al., 2024; Taylor et al., 2024; Xu et al., 2023), to assist providers’ delivery of psychotherapeutic interventions (Sharma et al., 2023), and as autonomous patient interaction agents (Sharma et al., 2024). The promise of LLMs, however, is predicated on the quality of the knowledge encoded within the models by the training process, and several studies have noted that the unfiltered base models can generate dangerous or harmful responses (Grabb et al., 2024). Successive models have shown improved performance on natural language tasks both because of increases in the size of their corpus (i.e. the volume of pre-existing text used to train the models) as well as the addition of additional human-level supervision, tuning, and model complexity.

The use of large-scale corpora collected from publicly available written and internet literature, however, may create limitations on the applicability of the model to the specialized tasks found in the practice of psychiatry and behavioral health. One study of user intention found that 78% of patient respondents were willing to use ChatGPT for self-diagnosis (Shahsavar and Choudhury, 2023), and in our experience, patients frequently use LLMs to evaluate their own mental health concerns, provide diagnostic and treatment recommendations, and even to provide autonomous psychotherapy, despite the potential risks of such usages and the stipulations of major vendors against using these tools for medical advice.

Few studies have directly examined the efficacy of LLMs on tasks related to knowledge or interpretation within behavioral health. Xu et al. (2023) developed and examined the efficacy of LLMs for predicting mental health-related metrics from Reddit posts, finding that LLMs were able to classify and stratify suicidality and depression from these short text snippets better than random. Galatzer-Levy et al. (2023) examined the ability of the *Med-PaLM 2* foundation model to analyze patient interviews and case vignettes and predict psychometric scores and diagnoses, finding that the model has promise in both applications. Here, we present a study investigating 1) the ability of the GPT family of LLMs (OpenAI, San Francisco, CA) to reason clinically about behavioral health and 2) the efficacy of integrating clinical expert-derived reasoning (through the use of decision trees) into the models to improve the accuracy of diagnostic prediction.

## 2. Methods

### 2.1 Dataset

The primary dataset for this study comprised case vignettes extracted from the DSM-5-TR Clinical Cases (Barnhill, 2023) book. Each example consisted of a multi-paragraph narrative vignette about a psychiatric clinical case and one or more DSM-5-TR (American Psychiatric Association, 2022) diagnoses assigned to the patient by the author of the vignette (“author-designated diagnoses”). The vignettes were organized by the DSM-5-TR diagnostic category corresponding to the patient’s primary diagnosis. A total of 106 cases were retrieved from the book. Cases from the sections on Elimination Disorders, Gender Dysphoria, Personality Disorders, and Paraphilic Disorders were discarded due to these DSM categories not being covered by the Handbook of Differential Diagnosis (see below). The remaining 93 cases were split into training and testing sets (of 38 and 55 cases respectively), using sampling stratified on primary diagnosis DSM category.

### 2.2 Inference approach and large language models

#### 2.2.1 Inference approach

Two inference approaches were implemented and compared for this study in order to evaluate the capability of the study LLMs to reason clinically about mental health and to integrate external expert reasoning into its predictions. In the first approach, termed the “base” approach, the LLM was directly prompted to assign diagnoses to the vignette, without the inclusion of outside knowledge or use of iterative prompting. In the second approach, termed the “decision tree” (DT) approach, a decision tree-based inference system was implemented. In this approach, the LLM is iteratively prompted with specific behavioral health questions regarding the input vignette. Candidate diagnoses are then predicted based on the answers to these questions, and then the list of candidate diagnoses is narrowed through additional queries to the LLM. Prompt templates for both approaches are shown in Table 1. The study was conducted between February and August 2024.

**Table 1.**
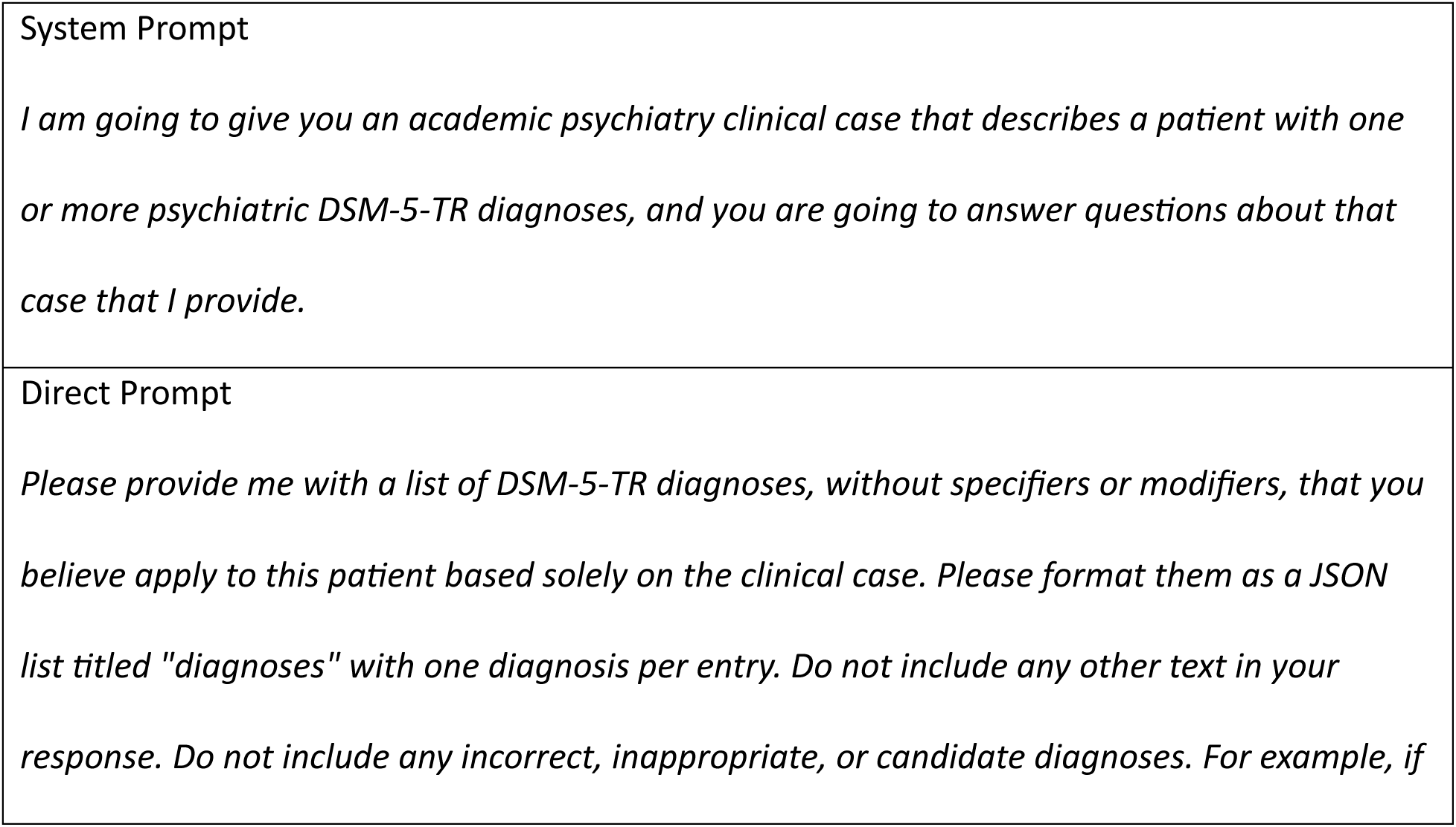
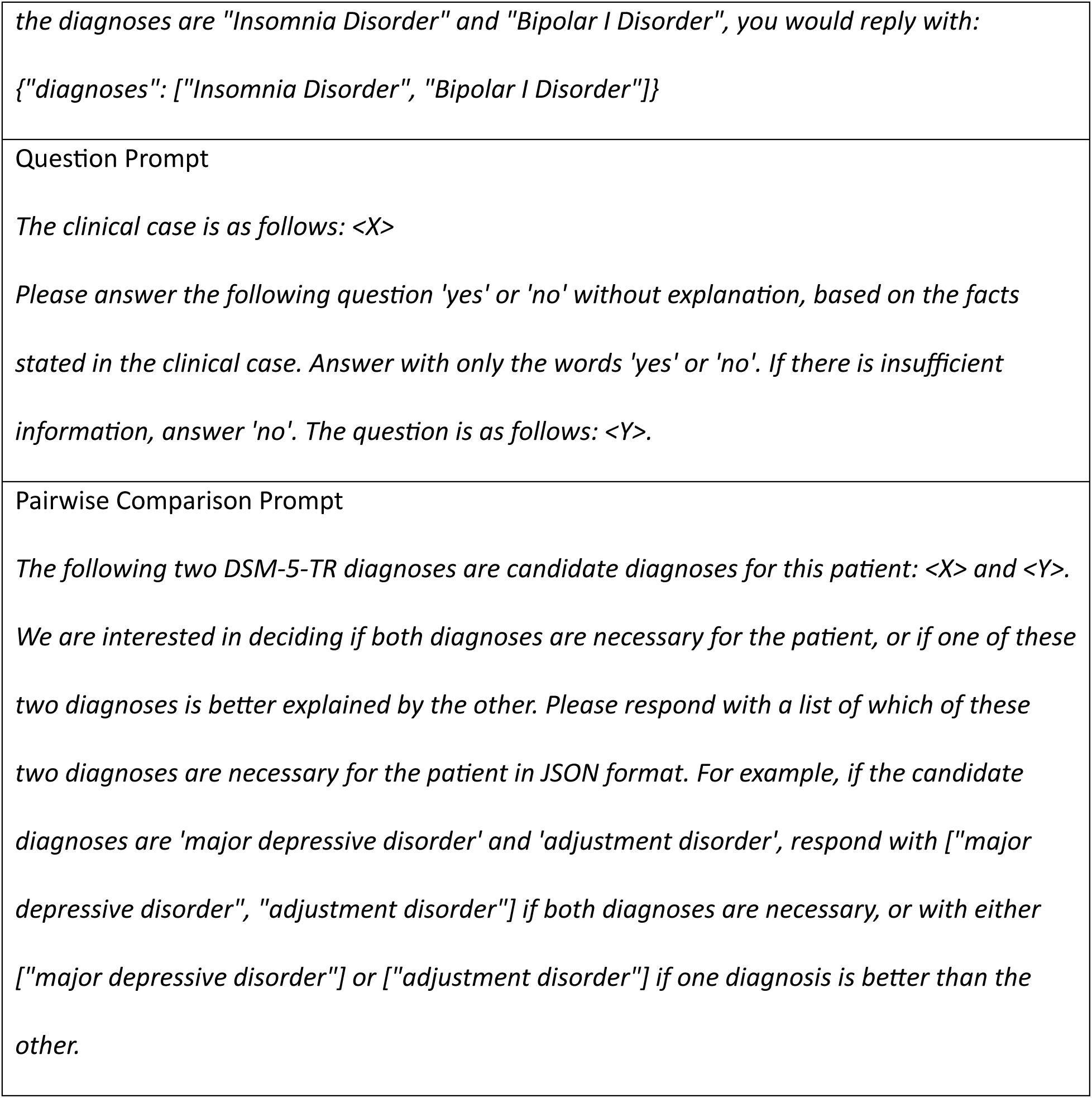
Prompt templates used for experiments.

#### 2.2.2 Decision tree implementation

The DSM-5-TR Handbook of Differential Diagnosis (the “Handbook”) was used as the expert knowledge source for the initial development of the decision tree prompting model (Michael B. First, 2024). This handbook consists of a series of 28 symptom-based decision trees for diagnosis. Each tree consists of a series of yes or no questions, the answers to which lead either to other questions or to diagnoses. All 28 trees were extracted and implemented as iterated yes or no prompts (“question prompts”). For each tree, a one-paragraph prompt was written describing the symptom category pertaining to the tree and asking if the vignette describes a patient experiencing that category of symptoms (“screening prompts”). See Table 1 for prompt templates and Appendix A for examples.

To make predictions using the decision tree model, each vignette was processed through each of the 28 decision trees. First, the screening prompt for each tree was used to determine which decision trees could be applicable to the vignette, and then for each applicable tree, the question prompts were used iteratively, collecting diagnoses from each tree into a list of candidate diagnoses for the vignette. To narrow the list of candidate diagnoses into the final list of diagnoses (“model-predicted diagnoses”), the model was prompted (see Table 1 for template) to compare each pair of candidate diagnoses and determine if both diagnoses were necessary, or if only one of them was needed.

#### 2.2.3 Large language models and parameters

For comparison, three successive versions of the commonly used *GPT* family of LLMs (OpenAI, San Francisco, CA) were evaluated: GPT-3.5, GPT-4, and GPT-4o, the “foundation models.” To ensure that the models did not learn from experimental inputs during the study, all queries were executed under an agreement to not use inputs, outputs, or any other data generated during the study for training purposes. Each query to the model was performed independently, without including context from other queries. Additional technical detail is available in Appendix B.

### 2.3 Decision tree refinement

In the first phase of the project, the decision tree models from the Handbook were refined through experimentation on the test set. The model was initially implemented using the exact decision trees provided in the source handbook. Then, the model was used to generate initial predictions on the testing cases. During this process, each prompt and the LLM response to the prompt was logged for review. These responses were then reviewed for every vignette. Based on this review, common themes of incorrect inferences based on the decision trees were developed, and the trees, screening prompts, and question prompts were then refined based on these common themes. The primary refinement approach was to address the discovered common themes through the use of known best practices for LLM prompt optimization, such as task decomposition and sequential tasking (Zhou et al., 2023) (dividing a complex task into smaller, specific tasks that are executed sequentially; e.g., breaking “Is the patient experiencing a Manic Episode” into a series of criterion-specific questions).

### 2.4 Diagnosis matching and simplification

To improve the initial tractability of the task, facilitate ease of comparison, and match the task most closely to the diagnostic power of the decision trees from the Handbook (which makes several simplifications to DSM-base diagnoses), all diagnoses (including author-designated and model-predicted) were systematically simplified using the procedure below:

1. All DSM specifiers, modifiers and codes were removed; if after this step diagnoses were identical, they were combined.
2. All neurocognitive disorders were collapsed into “Delirium” or “Neurocognitive Disorder,” removing disease-specific language and combining the mild and major classes.
3. All substance-specific disorders were collapsed into “Substance Use Disorder,” “Substance Intoxication,” or “Substance Withdrawal” (removing identification of the specific substance)
4. All breathing-related sleep disorder diagnoses (as defined by the DSM Sleep-Wake Disorders section) were combined into a single “Breathing-Related Sleep Disorder” diagnosis
5. All “Other Specified” and “Unspecified” diagnoses were combined into single “Other Specified or Unspecified” diagnoses for each category

After prediction and simplification, all base model-generated diagnoses further underwent a matching process to associate it with an exact DSM-5-TR diagnosis when possible as follows:

1. If the diagnosis exactly matched a DSM-5-TR diagnosis, this diagnosis was used.
2. If the diagnosis was clearly non-psychiatric (e.g., “Hypertension”) or was clearly related to a Z code (e.g., “Nonsuicidal Self-injury”), it was discarded.
3. If the diagnosis was psychiatric and had a clear, unambiguously matching DSM-5-TR diagnosis, it was replaced with the DSM-5-TR diagnosis (e.g., “Major Depression Disorder” to “Major Depressive Disorder”).
4. If there was no clear, unambiguously matching DSM-5-TR diagnosis, either because the diagnosis was not/no longer in the DSM-5-TR (e.g., “Sexual Aversion Disorder,” “Postpartum Psychosis,” or “Bullying Victimization”), or because the diagnosis was underspecified (e.g., “Bipolar Disorder”), the diagnosis was retained (and assigned as a false positive).

Each diagnosis was then also assigned a category based on the DSM-5-TR category in which the diagnosis is found (e.g., “Depressive Disorders”). Unmatched diagnoses were assigned a relevant category if a clear, unambiguously matching category was found (e.g., “Bipolar Disorder” to “Bipolar and Related Disorders”), or otherwise were assigned the category “Non-DSM.”

### 2.5 Analysis

For each vignette, base and DT model-generated inferences were created for each foundation model and then compared to the vignette author-designated diagnoses. Model-generated diagnoses were scored as a True Positive (TP) if they matched one of the author-designated diagnoses, or otherwise as a False Positive (FP). Author-designated diagnoses were scored as a False Negative (FN) if there was no matching model-generated diagnosis. This scoring approach was also used for associated categories. After scoring, the positive predictive value (PPV) and sensitivity were calculated for each testing set vignette, the composite F_1_ performance score was calculated from these scores, and then all were averaged across the dataset for reporting (see Appendix C for equations); in circumstances where the calculation would cause a division by zero, 0 was substituted for the result. For statistical testing, the paired Student’s *t*-test was used. For each foundation model, a two-tailed test was performed to compare PPV, sensitivity and F_1_ between the Base and DT models using a significance level of 0.05.

## 3. Results

### 3.1 Decision tree refinement

Decision tree refinement was carried out using the 38 training set vignettes. Nine categories for refinement of the Handbook trees were identified from the initial experiment; eight of which were based on disorder groupings and one of which was used for changes to the overall tree structure. For each category, a set of refinements was developed and implemented into the decision tree model prompting system. Common refinements included expanding definitions of specialized words used in behavioral health (e.g., “egosyntonic”) and expanding references to criteria to include the full criteria (e.g., prompting specifically for each of the criteria of a Manic Episode rather than prompting the model to determine if “criteria for a Manic Episode are met”). A summary of the categories and implemented refinements is presented in Table 2.

**Table 2.** Result and classification counts for predicted diagnoses across experiments. Numbers reflect results after post-processing. FM: Foundation Model, TP: True Positives, FP: False Positives, FN: False Negatives, DT: Decision Tree Model.

|  | FM | Model | Author Dx | Model Dx | TP | FP | FN |
| --- | --- | --- | --- | --- | --- | --- | --- |
| Diagnosis | gpt-3.5 | Base | 77 | 163 | 55 | 108 | 22 |
|  |  | DT | 77 | 76 | 46 | 30 | 31 |
|  | gpt-4 | Base | 77 | 163 | 61 | 102 | 16 |
|  |  | DT | 77 | 96 | 54 | 42 | 23 |
|  | gpt-4o | Base | 77 | 161 | 62 | 99 | 15 |
|  |  | DT | 77 | 89 | 55 | 34 | 22 |
| Category | gpt-3.5 | Base | 73 | 133 | 68 | 65 | 5 |
|  |  | DT | 73 | 67 | 53 | 14 | 20 |
|  | gpt-4 | Base | 73 | 135 | 70 | 65 | 3 |
|  |  | DT | 73 | 82 | 62 | 20 | 11 |
|  | gpt-4o | Base | 73 | 134 | 70 | 64 | 3 |
|  |  | DT | 73 | 80 | 62 | 18 | 11 |

### 3.2 Inferences, diagnosis matching and simplification

Inferencing, diagnosis matching, and simplification were carried out on the 55 test set vignettes. There were a total of 77 author-designated diagnoses and 73 author-designated categories across the test set vignettes. After completing diagnosis matching, a small number of base model diagnoses were not matchable to a DSM-5-TR diagnosis, a DSM-5-TR *Z* code, or a non-psychiatric illness; three such diagnoses were noted for *gpt-3.5*, one for *gpt-4*, and three for *gpt-4o*. This represented less than 1% of the total number of predicted diagnoses for each model. At the diagnosis level of specificity, the Base models predicted 161-163 diagnoses, with TP, FP, and FN ranges of 55-61, 99-108, and 15-22 respectively; the DT models predicted 76-96 diagnoses, with TP, FP, and FN ranges of 46-55, 30-42, and 22-31 respectively. At the category level of specificity, the Base models predicted 133-135 categories, with TP, FP, and FN ranges of 68-70, 64-65, and 3-5, respectively; the DT models predicted 67-82 categories, with 53-62, 14-20, and 11-20, respectively. Diagnosis and category counts and classifications after analysis are found in Table 3.

**Table 3.** Results of performance analysis of predictions for diagnosis and diagnostic category, by experiment. Bold text and * denote values that are statistically significantly higher by paired two-tailed t-testing between Base and DT models. FM: Foundation Model, PPV: Positive Predictive Value, DT: Decision Tree Model.

|  |  | By Diagnosis |  |  | By Category |  |  |
| --- | --- | --- | --- | --- | --- | --- | --- |
| FM | Model | Sensitivity | PPV | F<sub>1</sub> | Sensitivity | PPV | F<sub>1</sub> |
| gpt-3.5 | Base | 68.33% | 35.42% | 0.4418 | **91.21%\*** | 56.97% | 0.6647 |
|  | DT | 59.09% | **60.30%\*** | **0.5703\*** | 75.15% | **80.91%\*** | 0.7539 |
| gpt-4 | Base | 77.27% | 42.12% | 0.5138 | **95.76%\*** | 58.33% | 0.6842 |
|  | DT | 70.91% | **60.73%\*** | **0.6315\*** | 83.33% | **75.15%\*** | **0.7709\*** |
| gpt-4o | Base | 77.58% | 43.27% | 0.5254 | 93.94% | 59.24% | 0.6870 |
|  | DT | 71.82% | **65.27%\*** | **0.6645\*** | 84.24% | **79.09%\*** | **0.7915\*** |

### 3.3 Statistical analysis

Analysis results by foundation model (FM), diagnosis model (Base or DT), and level of granularity (diagnosis or category) are shown in Table 4. At the diagnosis level of specificity, the DT model had significantly higher PPV and F_1_ scores than the base model (with an average increase of +22%, +0.13, respectively) for all FMs, without a finding of significance for comparisons of the sensitivity. At the category level of specificity, the DT model had significantly higher PPV (average +20%) for all FMs but had significantly lower sensitivity for the *gpt-3.5* and *gpt-4* FMs (average −13% across all 3 models), leading to a significant increase in F1 for the *gpt-4* and *gpt-4o* FMs (average +0.09 across all 3 models).

## 4. Discussion

In this paper, we sought to evaluate the capabilities of the *GPT* family of large language models when applied to psychiatric reasoning and diagnosis, and to evaluate whether directly integrating clinician-expert guidance (in the form of decision trees) into the models improved their psychiatric performance. To this end, we evaluated two paradigms for utilizing the LLMs to make diagnoses: 1) directly prompting the models to predict diagnoses without access to knowledge external to the model (the Base approach), and 2) adapting expert-created diagnostic decision trees into sequential prompts in order to produce candidate predictions, and then prompting the model to determine which candidates were most appropriate. Overall, we found that both approaches were able to make appropriate diagnostic predictions, with some trade-offs noted between the two approaches. This finding demonstrates that the underlying foundation models do have psychiatric reasoning capabilities, despite not being specifically trained for this use case.

### 4.1 How do LLMs perform when directly prompted to estimate diagnoses?

In direct prediction efforts using the Base approach, we found that the model was able to correctly produce the majority of correct diagnoses, with the sensitivity mean ranging from 68% (*gpt-3.5*) to 78% (*gpt-4o*). This is concordant with the findings of Galatzer-Levy et al. (2023), who used a different family of models in a restricted subset of vignettes and diagnoses and found 77.5% accuracy in predicting the correct primary diagnosis. When assessing whether the model predicted diagnoses in the correct DSM-5-TR category (rather than whether the specific diagnoses were correct), the Base approach demonstrated impressive sensitivity means of 91% (*gpt-3.*5) to 95% (*gpt-4*). We interpret this result as demonstrating that the foundation models have an inherent capacity to extract symptoms and mental health concerns from narratives and reason about likely diagnoses using this information; indeed, the degree of concordance between the predicted diagnoses and the author-designated diagnoses is superior to that found for most of the mental disorders studied in the DSM-5 field trials (Clarke et al., 2013; Regier et al., 2013) (which found pooled intraclass Kappa statistics of 0.46 for schizophrenia, 0.56 for bipolar disorder, and 0.28 for major depressive disorder).

However, we also found that fewer than half of the predicted diagnoses were correct, with a PPV mean ranging from 35% (*gpt-3.*5) to 43% (*gpt-4o*) for specific diagnoses and 57% to 59% respectively for diagnostic categories. We hypothesize that the significant overdiagnosis rates represent the model’s limited capability to apply criteria and clinical judgement to determine if a patient’s symptoms meet the DSM-5-TR defined threshold for a behavioral health diagnosis. The authors find the propensity of the LLMs for overdiagnosis to be particularly concerning given the empiric prevalence in our practice of patients who make use of publicly available LLMs for self-diagnosis; their usage of these systems is closest to our Base approach and is likely similarly subject to the same limitation. Clinicians should ensure their patients are adequately informed about this risk in the use of LLMs to assist patients in managing their own mental health. The degree of risk and potential harm may vary by patient, and future work could study how emergent patient-driven uses of public LLMs impact both the general population and people with mental disorders. Since it is likely that such uses will only increase as these technologies continue to permeate the public consciousness, the development of guidelines to prevent inappropriate clinical use of these technologies is of great importance.

### 4.2 Does the integration of expert decision trees improve the diagnostic capabilities of LLMs?

In decision tree-based prediction efforts using the DT approach, we found the model was again able to correctly produce the majority of correct diagnoses, with sensitivity means ranging from 59% (*gpt-3.5*) to 71% (*gpt-4o*); we did not find a statistically significant difference between the two approaches in sensitivity scores for specific diagnoses when using the paired Student’s *t*-test. When looking at diagnostic categories, we did find that for the *gpt-3.5* and *gpt-4* models, the DT approach had statistically significantly lower sensitivity scores than the Base approach. When examining PPV, however, we found that the DT approach demonstrated a statistically significant improvement for all models across specific diagnosis and diagnostic category. This led to a significant improvement in F_1_ score using the DT approach for all experiments except for the evaluation of performance in diagnostic category for the *gpt-3.5* model.

We hypothesize that these results demonstrate that the integration of the decision trees improved the capability of the model to apply diagnostic criteria in order to determine if behavioral health symptoms met the threshold of a diagnosis, leading to an improvement in PPV across all experiments. The concomitant reduction in sensitivity performance is consistent with the known tradeoff between the two metrics when increasing the “threshold” for predicting a positive class. This may also represent increasing difficulty in the model answering specific questions about diagnostic criteria from the unstructured vignette (as opposed to the easier task of extracting broad symptoms). Of note, we found that significant adaptation to the decision tree models found in the Handbook were required in order to optimize performance for our study. This is consistent with previously reported results (Nori et al., 2023; Zhou et al., 2023) that have demonstrated the impact of the application of prompt engineering techniques on model performance.

### 4.3 How has the progression of GPT models impacted their performance in psychiatric reasoning?

We found that, broadly, performance tended to improve with the use of successive *GPT* models. The biggest jump in performance by F_1_ score was between the *gpt-3.5* and *gpt-4* models for both approaches, with a more modest increase between *gpt-4* and *gpt-4o*. This is consistent with reported results in other domains(Shahriar et al., 2024), and may reflect in part that changes to the foundation models are aimed both to improve performance and to reduce the cost of inference – goals that often require tradeoffs. Notably, inference costs are significantly different between the three models. These costs are priced in units of 1M input and output tokens, and are $0.50/$1.50 for *gpt-3.5*, $10/$30 for *gpt-4*, and $5/$15 for *gpt-4o*.

#### What are the potential limitations of this work and opportunities for further investigation?

This effort represents the first comprehensive evaluation of the performance of *GPT* models in psychiatric diagnosis with and without the use of external expert guidance. Executing the study required two technical compromises that may represent potential limitations. In order to make use of standardized, commonly-available input examples that were not designed specifically for AI applications, case vignettes were obtained using the APA’s published casebook. Given that the *GPT* models were trained on corpora that include published books, it is possible that current or former versions of the casebook were included in the training dataset. This might have provided the models with an advantage in making diagnoses using these examples that would not be present when analyzing other data. We believe that the probability of significant influence on our results is low due to the length of the vignette examples. Additionally, there is no published material on the application of the Handbook decision trees on the casebook vignettes, reducing the probability that memorization would affect the DT results.

Additionally, in order to facilitate direct comparison between author-designated diagnoses and predicted diagnoses, manual review was used with a standardized diagnosis simplification system. This approach could have hidden important differences between the model’s output and the author-designated diagnoses. We believe that this potential trade-off was worthwhile given that our goal was to evaluate the model’s ability to reason about diagnosis and make use of expert guidelines, but future efforts may wish to investigate the performance of the models in more specific areas, such as the generation of appropriate DSM specifiers or in differentiation between types and severity of neurocognitive disorders or substance use disorders. Additionally, we excluded vignettes with primary diagnoses from DSM-5-TR chapters not covered by the Handbook, such as personality disorders and paraphilic disorders; future efforts could investigate the adaptation of other expert resources that apply to those areas.

More broadly, future efforts could investigate the use of LLMs for other types of psychiatric reasoning. DSM diagnosis has limited interrater reliability (Clarke et al., 2013; Regier et al., 2013), and in the absence of a human comparison arm, our ability to evaluate the significance of the model’s performance is restricted.

The advent of LLMs and other advanced artificial intelligence-based modeling tools could allow for the development of new diagnostic schema that could overcome some of the limitations of the DSM through the automated interpretation of large volumes of patient-related data. Such an approach could allow for more precise quantification of language-based phenotypes, motivate new approaches for disorder subtyping, and allow for the discovery of new links between behavioral phenotypes and neurobiological mechanisms, with the overall goal of matching the right treatments to the right patients at the right time.

### 4.4 Conclusion

In this work, we have demonstrated that the *GPT* family of large language models has the emergent capability for psychiatric reasoning and that it is able to interpret case vignettes and apply expert guidelines to make diagnoses. We found that directly prompting the models without external information led the models to predict the majority of correct diagnoses, with the limitation of significant overdiagnosis. We found that incorporating adapted expert decision-tree based diagnostic guidelines reduced overdiagnosis and improved overall model performance. These results illustrate the potential risks and benefits of the use of large language models for language analysis in behavioral health and motivate the need for systems that integrate language modeling with expert knowledge for use in clinical applications.

## Data Availability

All data produced in the present study are available upon reasonable request to the authors; underlying data is available from the primary sources from which it is derived

## Acknowledgements

We thank the UCSF AI Tiger Team, UCSF Academic Research Services, UCSF Research Information Technology, and the UCSF Chancellor’s Task Force for Generative AI for their support in developing LLM resources used for this project. This research was made possible through the use of content belonging to the American Psychiatric Association; express permission was obtained from the American Psychiatric Association for the use of such content (DSM-5-TR Clinical Cases and DSM-5-TR Handbook of Differential Diagnosis (Copyright © 2023 and 2024). American Psychiatric Association. All Rights Reserved, including rights for text and data mining (TDM), Artificial Intelligence (AI) training, and similar technologies).

## Appendices

### Appendix A. Decision Tree Prompting

For the initial experiments (prior to refinement), question prompts were created based on suggested language in the DSM-5-TR Handbook of Differential Diagnosis. For example, the following is an example from the “Depressed Mood” decision tree:

> *Are there at least 2 weeks of depressed mood or diminished interest plus associated characteristic symptoms (e.g., changes in weight and appetite, fatigue, feelings of worthlessness or guilt, changes in sleep, suicidal thoughts)?*

A screening prompt was developed for each decision tree, such as the following example from the “Self-Injurious Behavior” tree:

> *For the purposes of this discussion, self-injurious behavior refers to intentional self-inflicted acts to injury or mutilate one’s own body. This includes include cutting, burning, head banging, hair pulling, skin picking, self-biting, and hitting of various parts of one’s own body, but does not include socially or culturally sanctioned practices (such as piercing or artistic scarification). It does not include behavior intended to end one’s own life. Based on this definition, is there evidence in the clinical case that the patient has self-injurious behavior?*

### Appendix B. LLM Technical Specifications

For this paper, three specific commercial large language models developed by OpenAI (San Francisco, CA) were used. The models used were *gpt-3.5*, *gpt-4-turbo*, and *gpt-4o*. To enable reproducibility, specific model versions were used as follows: *gpt-3.5-turbo-0125*, *gpt-4-0125-preview,* and *gpt-4o-2024-05-13*. A python interface was used to execute queries to the foundation models using the OpenAI API. All configurable content filters were turned off, and all queries were made with a temperature of 0, *top_p* of 1, and without frequency or presence penalties. If a query returned either an error or an unparseable response, the query was repeated with exponential backoff until a valid response was received. All queries ultimately produced parseable responses using this method, and no queries were rejected due to content filtering.

### Appendix C. Performance Metric Equations

The following equations were used to calculate performance metrics:

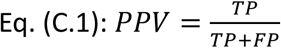

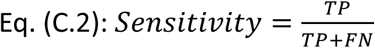

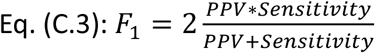

---

**Table 1.**
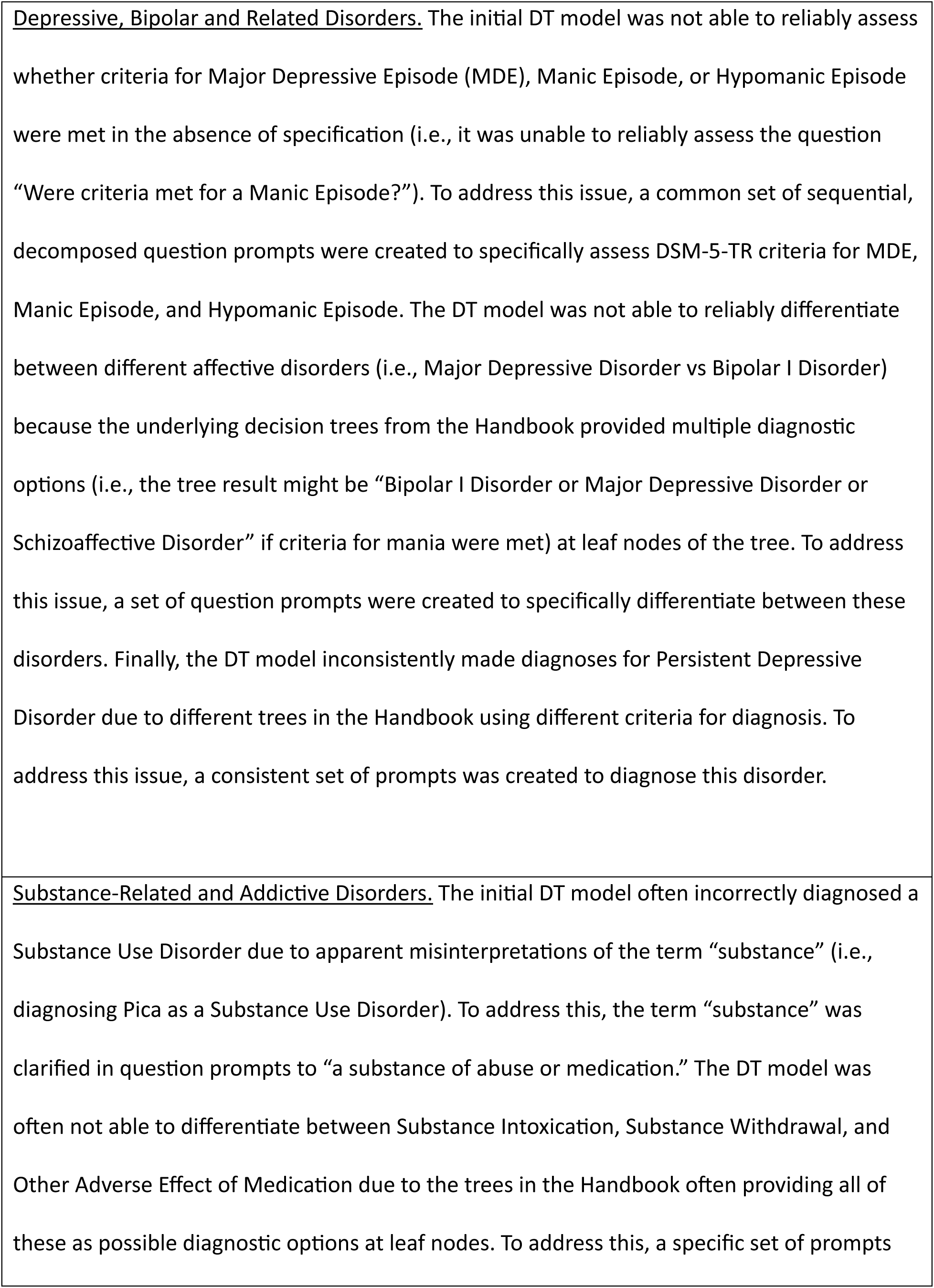
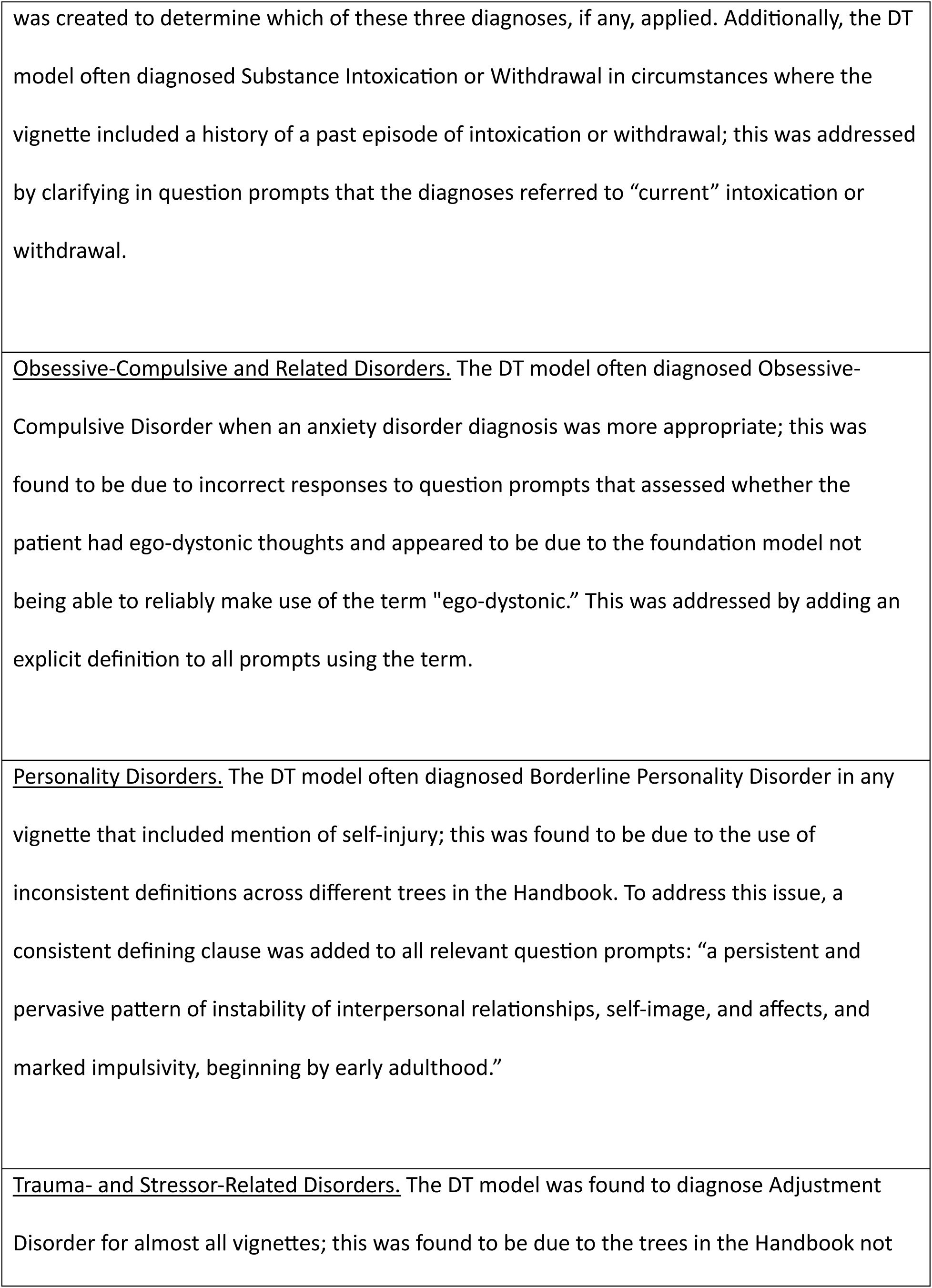
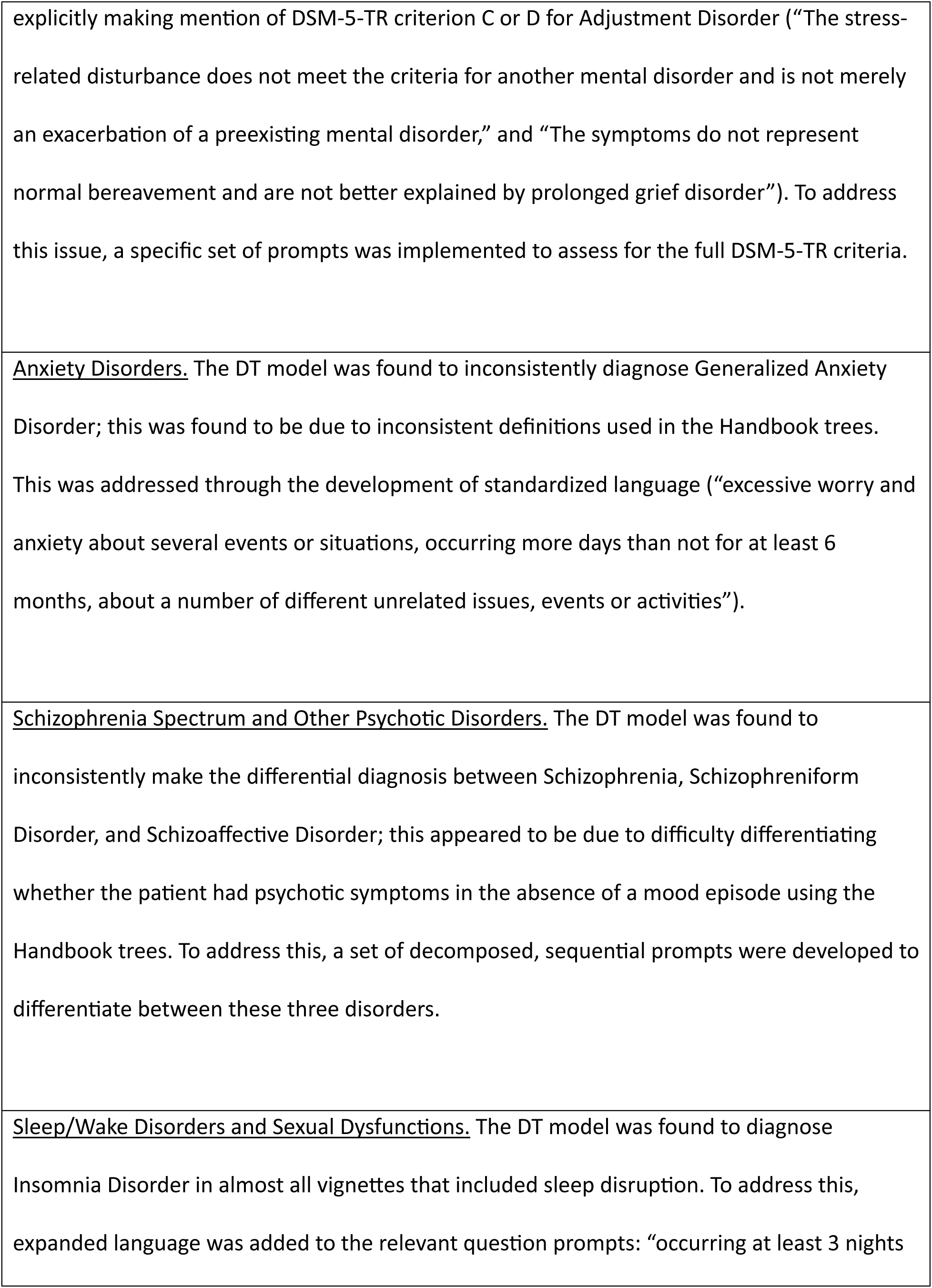
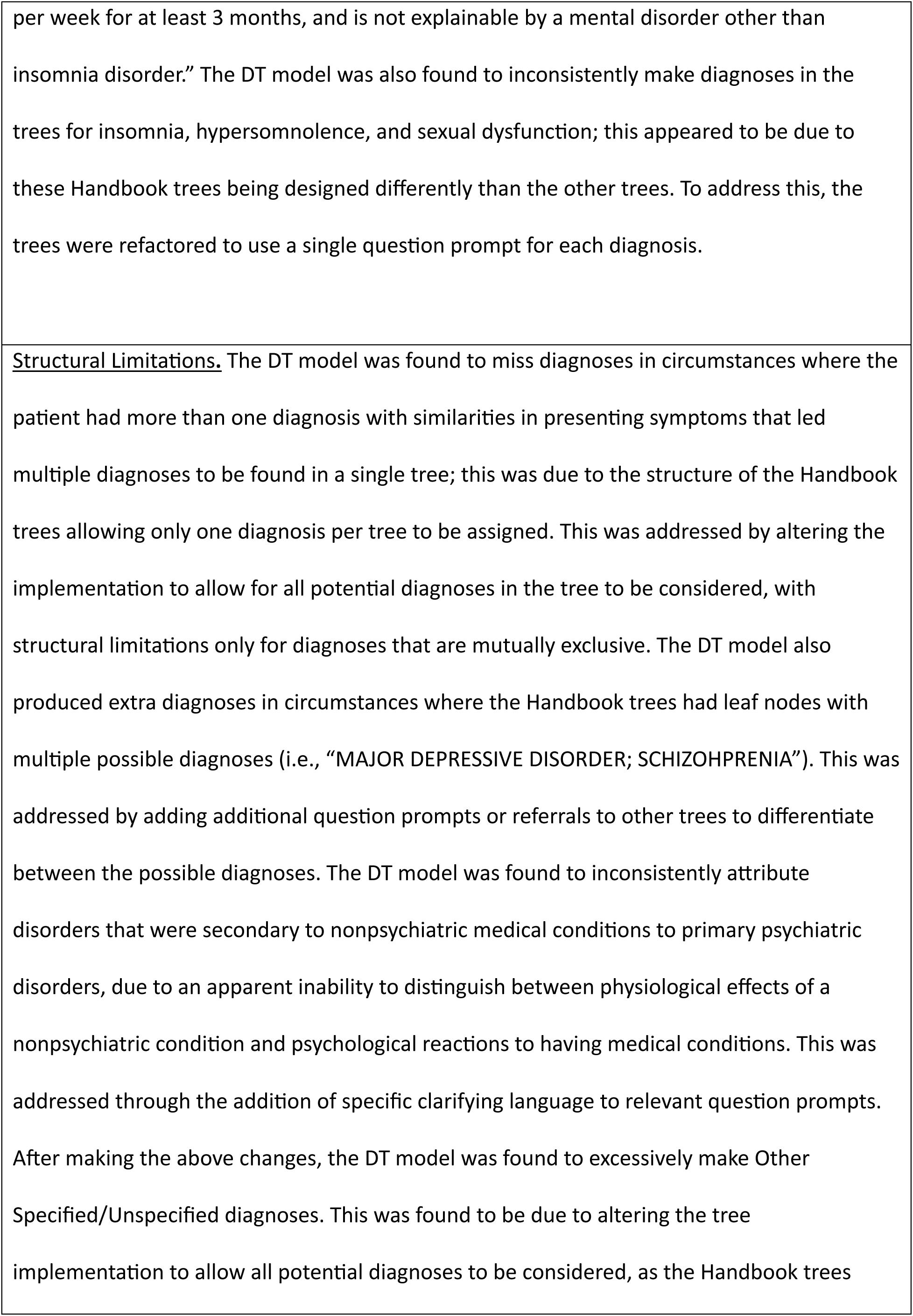
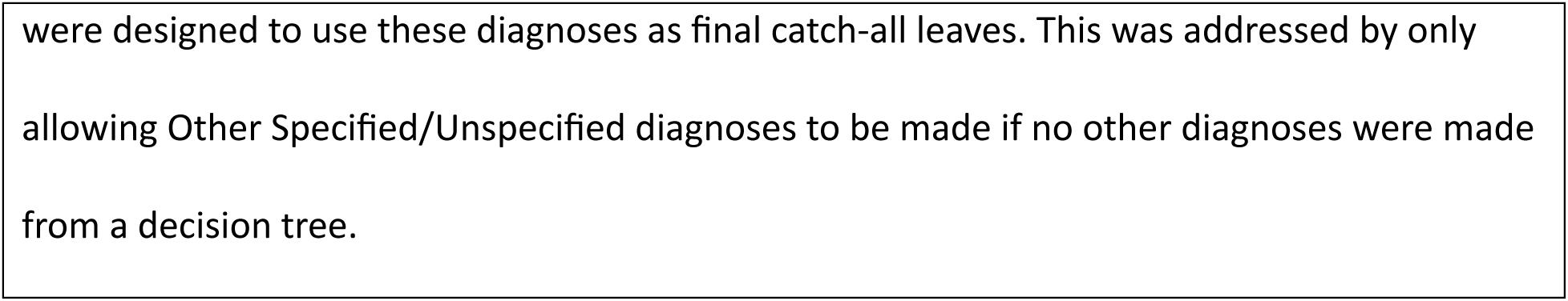
Findings and results of decision tree refinement process by category.

